# Early Treatment of Favipiravir in COVID-19 Patients Without Pneumonia: A Multicentre, Open-Labelled, Randomized Control Study

**DOI:** 10.1101/2022.06.06.22275902

**Authors:** Rujipas Sirijatuphat, Weerawat Manosuthi, Suvimol Niyomnaitham, Andrew Owen, Katherine K. Copeland, Lantharita Charoenpong, Manoch Rattanasompattikul, Surakameth Mahasirimongkol, Kulkanya Chokephaibulkit

## Abstract

We investigated Favipiravir (FPV) efficacy in mild cases of COVID-19 without pneumonia and its effects towards viral clearance, clinical condition, and risk of COVID-19 pneumonia development. PCR-confirmed SARS-CoV-2-infected patients without pneumonia were enrolled (2:1) within 10 days of symptomatic onset into FPV and control arms. The former received 1800 mg FPV twice-daily (BID) on Day 1 and 800 mg BID 5-14 days thereafter until negative viral detection, while the latter received supportive care only. The primary endpoint was time to clinical improvement, which was defined by a reduced National Early Warning Score (NEWS) or score of ≤1. 62 patients (41 female) comprised the FPV arm (median age: 32 years, median BMI: 22 kg/m²) and 31 patients (19 female) comprised the control arm (median age: 28 years, median BMI: 22 kg/m². The median time to sustained clinical improvement by NEWS was 2 vs 14 days for FPV and control arms respectively (adjusted hazard ratio (aHR) of 2.77, 95% CI 1.57-4.88, *P* <0.001). The FPV arm also had significantly higher likelihoods of clinical improvement within 14 days after enrolment by NEWS (79% vs 32% respectively, *P* <0.001), particularly female patients (aOR 6.35, 95% CI 1.49-27.07, *P* <0.001). 8 (12.9%) and 7 (22.6%) patients in FPV and control arms developed mild pneumonia at a median (range) 6.5 (1-13) and 7 (1-13) days after treatment, respectively (*P* = 0.316); all recovered well without complications. We can conclude that early treatment of FPV in symptomatic COVID-19 patients without pneumonia was associated with faster clinical improvement.

## Introduction

Across 226 countries and territories, over 517 million cases and 6.2 million deaths have been recorded for SARS-CoV-2 as of May 13^th^, 2022 [1]. Its viral spread relies on angiotensin converting enzyme 2 (ACE-2) receptor binding, RNA-dependent RNA polymerase (RdRp), as well as other host and viral proteins important for successful transmission and replication [2,3]. In 80-90% of these cases, the infection is self-limiting and relatively mild or moderate, bearing a presentation and organ tropism similar to Influenza [2-5]. However, some patients experience life-threatening complications and post-acute COVID-19 syndrome [2,6-10]. Several interventions have emerged in the past 2 years for treating COVID-19, but there remains a critical need for widely available medicines. Accordingly, multiple existing antivirals are being investigated for their suitability to be repurposed and studied as possible treatment options [2,3,11-14].

Easily utilizable antivirals capable of inhibiting SARS-CoV-2 replication mechanisms are greatly desired – particularly for treating mild-to-moderate cases, which comprise a majority of reported illnesses. Such treatments could help prevent downstream complications and diminish transmission. Remdesivir (RDV) was one such early-implemented, USFDA-approved antiviral [15]. It had been used in multiple clinical trials to treat moderate or severe cases of COVID-19 with pneumonia or under oxygen supplementation [11,16-18], and in non-hospitalized patients [19]. Its limited clinical application, high cost ($390 per 100 mg vial), and intravenous mode of administration rendered RDV less applicable in resource-limited countries afflicted by a large number of COVID-19 cases [11,17]. In addition to RDV, molnupiravir (MPV) and nirmatrelvir/ritonavir have also recently received conditional approvals in some territories for use in outpatients. Further to publication of phase II/III MOVe-OUT trial for MPV [20], several commentators have however queried the available data [21,22] and larger studies are ongoing to clarify the utility in different patient groups.

Favipiravir (FPV) is another promising antiviral drug. It is a broad-spectrum antiviral previously used to treat numerous diseases, including re-emerging or novel cases of Influenza [23,24], and has since been repurposed to treat SARS-CoV-2 [3,5,25]. FPV was shown to inhibit SARS-CoV-2 *in-vitro* in infected Vero E6 cells [11,12,26] and appeared to improve clinical outcomes, control viral progression, and promote viral clearance in numerous clinical studies [3,4,6,8,9,27,28]; however, the clinical benefit was not clearly demonstrated in some studies [29-33]. More than 4000 well-characterized patient safety profiles illustrated that effective FPV drug concentrations remained within safe therapeutic dosages [6,26,34]. This, coupled with it being easily, orally administrable (200 mg/tablet of AVIGAN) [35] and having a relatively low cost ($0.5-1.0 per pill) compared to RDV, MPV, and nirmatrelvir/ritonavir render it a worthy candidate for further evaluation [17,36]. Some countries have already approved, even commercialized, its use for treating mild or moderate COVID-19 [6,31,37-41].

RDV, MPV and FPV are all nucleoside-based drugs, which target the viral RdRp. However, unlike RDV that exerts its antiviral action by chain termination, MPV and FPV elicit their effects via a mechanism termed lethal mutagenesis [42,43] In this mechanism, the active metabolite is incorporated into the genomic or sub-genomic RNA, rather than endogenous nucleosides while copying the RNA template genome. The resultant drug-containing RNAs are then themselves used as a template for production of subsequent RNAs causing mistakes to be made in copying, to an extent that mutated genomes are not thought to form functional viruses.

Despite limited evidence of its benefits, Thailand has deployed FPV to treat COVID-19 since the start of the pandemic. While initial studies showed promise, particularly upon early treatment (<4 days after infection) [44,45], more clinical trials are required to further support and characterize FPV’s clinical applications [46,47]. In this study, we sought to investigate the efficacy of early FPV treatment towards clinical benefit, viral clearance, and risk of developing COVID-19 pneumonia in mild cases of COVID-19 without pneumonia.

## Methods

This multicentre, open-labelled, randomized prospective cohort took place from December 2020 to July 2021 at three medical centres in Bangkok: Bamrasnaradura Infectious Diseases Institute, Golden Jubilee Medical Centre, and Faculty of Medicine Siriraj Hospital. Eligible subjects were PCR-confirmed SARS-CoV-2 infected individuals, 18 years or older, with mild-to-moderate symptoms and without pneumonia. Subjects with pneumonia, in critical condition, that had a symptomatic onset >10 days, were suspected or confirmed to have concurrent or concomitant infections, that received immunosuppressive treatment, received or were on medication with possible SARS-CoV-2 antiviral activity (*e*.*g*., interferon alpha, lopinavir, chloroquine, hydroxychloroquine, ivermectin, favipiravir, and remdesivir), pregnant or possibly pregnant, or lactating were excluded from the study. After providing written informed consent, participants were randomized 2:1 into FPV and control arms. On top of supportive care, the FPV arm received oral administrations of FPV (Fujifilm Toyama Chemical co., Ltd.; 200 mg per tablet) as a dosing regimen of 1800 mg twice-daily (BID) for 1 day (9 tablets per dose) and 800 mg BID (4 tablets per dose) thereafter until clinical improvement or saliva RT-PCR became negative (min-max of 5-14 days). The control arm received only symptomatic therapy. This study was approved by each study site’s institutional ethic committee and was conducted in accordance with the Declaration of Helsinki, Belmont Report, and International Council on Harmonisation’s Good Clinical Practice. The study was registered in thaiclinicaltrials.org (TCTR20200514001) [48].

Patients were hospitalized for at least 7 days. Clinical findings, clinical symptoms, and oxygen saturation (SpO_2_) were reported daily, and vital signs were recorded twice-daily during hospitalization. Laboratory tests for monitoring safety (included haematology and chemistry) were performed on days 1, 4, 7, 10, 15, 22, and 28; saliva SARS-CoV-2 RNA (viral load) and chest imaging were performed every 3 days from days 1-28; and 12-lead ECG values were recorded on days 1, 14, and 28.

Efficacy evaluation included: the duration and resolution of pyrexia (body temperature ≤ 37.4 °C); clinical severity assessed by National Early Warning Score (NEWS) that based on individual physiological parameters (respiration rate, SpO_2_, any supplemental oxygen, temperature, systolic blood pressure, heart rate, and level of consciousness) [49]; absence of new chest imaging findings; and negative SARS-CoV-2 RT-PCR result from saliva specimen.

### Statistical Analysis

Clinical improvement was defined as reduced NEWS from baseline or score of ≤1. Sustained clinical improvement was defined as an improvement for 7 days or until discharge. All endpoints were monitored from the start of FPV administration to 28 days.

Descriptive analyses were conducted to provide general information about the patients in each arm, where continuous variables were reported as mean, standard deviation (SD), median, interquartile range (IQR), and categorical variables were reported as absolute (numbers) and relative (percentage) frequencies.

Log-rank tests were performed to test whether there was a difference in probability between patient arms from treatment initiation to ‘sustained improvement’ by NEWS. Primary endpoints were right-censored on Day 28, due to late sustained improvement of patients in the study. Graphs were generated using the Kaplan-Meier method, presenting survival probability as confidence intervals. Hazard ratios were derived using the regression coefficient of the Cox model – adjusted for demographic characteristics and SARS-CoV-2 viral load level as covariates. Proportional hazards (PH) assumption was checked using statistical tests and graphical diagnostics based on scaled Schoenfeld residuals.

Viral load clearance rate was determined by comparing the cumulative percent viral load level that fell below the detection limit and were compared within group using paired *t* test. Graphs were generated using the Kaplan-Meier method. Binary logistic regression was conducted to identify factors associated with sustained improvement within the first 14 days after receiving treatment. Models were checked for collinearity and homoscedasticity, and robust standard errors were used. Data was analysed using STATA, version 15.1 (STATA Corp, College Station, TX, USA), with a statistical significance of *P* < 0.05). Figures were visualized using GraphPad Prism 9.0 software and R program.

## Results

Of the 93 participants enrolled, 62 patients (41 female) comprised the FPV arm, with a median age of 32 years (IQR of 27-39 years), median BMI of 22 kg/m² (IQR of 19-25 kg/m²); and 31 patients (19 female) comprised the control arm, median age of 28 years (IQR of 25-35 years), median BMI of 22 kg/m² (IQR of 19-26 kg/m²) (Table 1). There was no significant difference in prevalence of underlying conditions (9.7% vs 6.5%), duration of COVID-19 symptoms before enrolment (mean 1.66 vs 1.64 days; 90% and 84% were <4 days respectively), and clinical presentations observed between the two arms – with the exception being the higher prevalence of fever in the FPV arm (29% vs 10%, Table 1). SARS-CoV-2 genotypes were similarly distributed between arms, with alpha being the predominant variant.

**Table 1.** Baseline characteristics and clinical symptoms of patients with SARS-CoV-2 infection.

| Variables | Favipiravir<br>(n=62) | Control<br>(n=31) | p-value |
| --- | --- | --- | --- |
| **Baseline characteristics** |  |  |  |
| Site; n (%) |  |  |  |
| Bamrasnaradura Infectious Diseases Institute | 51 (82.3) | 26 (83.9) | 1.000 |
| Golden Jubilee Medical Centre | 9 (14.5) | 4 (12.9) |  |
| Siriraj Hospital | 2 (3.2) | 1 (3.2) |  |
| Age; Median (IQR) | 32 (27-39) | 28 (25-35) | 0.044 |
| Gender; n (%) |  |  |  |
| Male | 21 (33.9) | 12 (38.7) | 0.646 |
| Female | 41 (66.1) | 19 (61.3) |  |
| **Baseline clinical symptoms** |  |  |  |
| Coughing | (n=61) | (n=30) |  |
| None | 14 (23.0) | 10 (33.3) | 0.156 |
| Mild | 41 (67.2) | 20 (66.7) |  |
| Moderate | 6 (9.8) | - |  |
| Sore throat | (n=61) | (n=30) |  |
| None | 22 (36.1) | 16 (53.3) | 0.178 |
| Mild | 35 (57.4) | 14 (46.7) |  |
| Moderate | 4 (6.6) | - |  |
| Headache | (n=61) | (n=30) |  |
| None | 46 (75.4) | 24 (80.0) | 0.862 |

| **Variables** | **Favipiravir**<br>**(n=62)** | **Control**<br>**(n=31)** | **p-value** |
| --- | --- | --- | --- |
| Mild | 14 (23.0) | 6 (20.0) |  |
| Moderate | 1 (1.6) | - |  |
| Muscle or joint pain | (n=61) | (n=30) |  |
| None | 48 (78.7) | 24 (80.0) | 1.000 |
| Mild | 12 (19.7) | 6 (20.0) |  |
| Moderate | 1 (1.6) | - |  |
| Nasal congestion or nasal discharge | (n=61) | (n=30) |  |
| None | 46 (75.4) | 26 (86.7) | 0.478 |
| Mild | 12 (19.7) | 4 (13.3) |  |
| Moderate | 3 (4.9) | - |  |
| Chills or sweating | (n=61) | (n=30) |  |
| None | 59 (96.7) | 30 (100.0) | 1.000 |
| Mild | 2 (3.3) | - |  |
| Moderate | - | - |  |
| Malaise or fatigue | (n=60) | (n=30) |  |
| None | 49 (81.7) | 28 (93.3) | 0.206 |
| Mild | 11 (18.3) | 2 (6.7) |  |
| Moderate | - | - |  |
| Diarrhoea | (n=22) | (n=12) |  |
| None | 17 (77.3) | 9 (75.0) | 1.000 |
| N/A | 5 (22.7) | 3 (25.0) |  |
| Loss of taste | (n=22) | (n=12) |  |
| No | 17 (77.3) | 8 (66.7) | 0.687 |
| N/A | 5 (22.7) | 4 (33.3) |  |
| Loss of smell | (n=22) | (n=12) |  |
| No | 13 (59.1) | 8 (66.7) | 0.881 |
| Yes | 4 (18.2) | 1 (8.3) |  |
| N/A | 5 (22.7) | 3 (25.0) |  |
| Fever |  |  |  |
| No | 44 (71.0) | 28 (90.0) | 0.035 |
| Yes | 18 (29.0) | 3 (10.0) |  |
| **Health conditions** |  |  |  |
| Body Mass Index (BMI: kg/m<sup>2</sup>); Median (IQR) | 22 (19-25) | 22 (19-26) | 0.8624 |
| Underweight (< 18.5 kg/m<sup>2</sup>) | 15 (24.2) | 7 (22.6) |  |
| Normal weight (18.5 - 24.9 kg/m<sup>2</sup>) | 30 (48.4) | 15 (48.4) |  |
| Overweight and obesity (> 24.9 kg/m<sup>2</sup>) | 17 (27.4) | 9 (29.0) |  |
| Have underlying health conditions |  |  |  |
| Yes | 6 (9.7) | 2 (6.5) | 0.601 |
| No | 56 (90.3) | 29 (93.5) |  |
| SARS-CoV-2 genotypes |  |  |  |
| Ancestral strain with D614G | 15 (26.8) | 5 (17.3) | 0.580 |
| B.1.1.7 (Alpha) | 37 (66.1) | 21 (72.4) |  |
| B.1.617.2 (Delta) | 4 (7.1) | 3 (10.3) |  |
| Duration of symptoms before treatment (day(s)); |  |  |  |
| Mean (SE) | 1.66 (2.4) | 1.64 (2.1) | 0.9748 |
| Median (IQR) | 0 (0-7) | 0 (0-6) |  |
| 0-4 days | 56 (90.3) | 26 (83.9) |  |
| > 4 days | 6 (9.7) | 5 (16.1) |  |
| NEWS Score |  |  |  |
| 0 | 24 (38.7) | 10 (32.2) | 0.648 |
| 1 | 24 (38.7) | 16 (51.6) |  |
| 2 | 10 (16.1) | 3 (9.7) |  |
| 3 | 4 (6.5) | 2 (6.5) |  |

The median time to sustained clinical improvement by NEWS was 2 days vs 14 (range of 1-28 days for both) for FPV and control arms respectively (adjusted hazard ratio (aHR) of 2.77, 95% CI 1.57-4.88, *P* <0.001) (Figure 1). Patients that received FPV also had significantly higher likelihoods of clinical improvement within 14 days after enrolment by NEWS (79% vs 32% respectively, *P* <0.001) (Figure 2). However, the proportion of patients with reported resolution of symptoms (*e*.*g*., dry cough, sore throat, headache, and nasal congestion) were not significantly different between arms (see Supplementary Figure 1).

**Figure 1.**
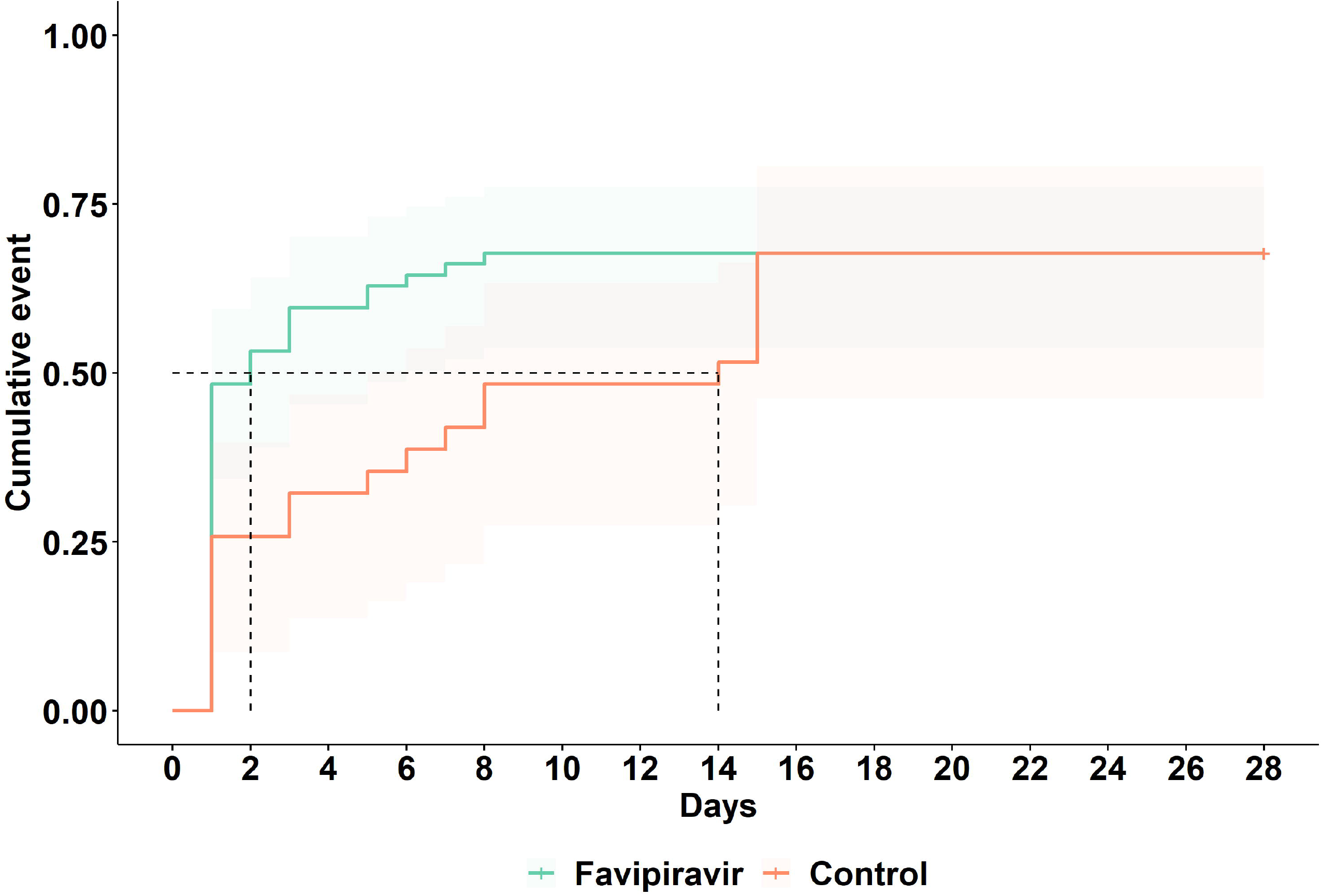
Time to sustained clinical improvement by NEWS. The Kaplan-Meier curve illustrates the cumulative proportion of patients who experienced sustained clinical improvement, which is defined by a reduced NEWS or NEWS ≤1 for at least 7 days. The median time to sustained clinical improvement by NEWS was 2 days vs 14 days (range of 1-28 days) for FPV and control arms respectively (adjusted hazard ratio (aHR) 2.77, 95% CI 1.57-4.88, *P* < 0.001)

**Figure 2.**
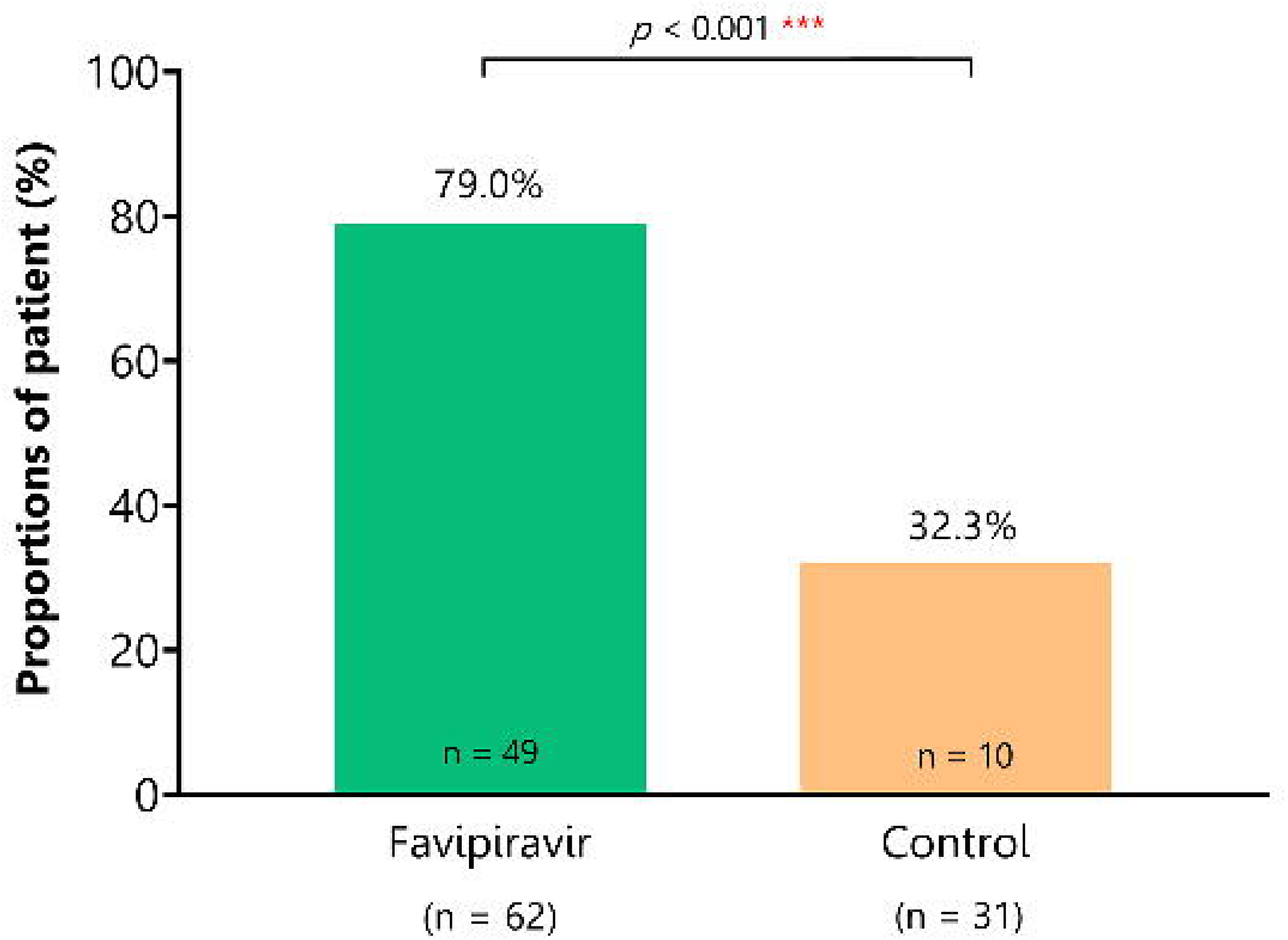
Proportion of patients with clinical improvement by NEWS within 14 days of treatment. The bar graph illustrates the cumulative proportion of patients who experienced clinical improvement, which is defined as reduced NEWS or NEWS ≤1 during the 14-day treatment period. Patients that received FPV also had significantly higher likelihoods of clinical improvement within 14 days after enrolment (79% vs 32% respectively, *P* < 0.001)

Among subjects in the FPV arm, female patients were more likely to experience clinical improvement within 14 days compared to male patients (aOR 6.35, 95% CI 1.49-27.07, *P* <0.001 (Table 2). Upon performing a regression analysis, FPV administration was the only independent factor associated with clinical improvement by NEWS within 14 days.

**Table 2.** Logistic regression analysis of associated factors with improvement of clinical conditions by NEWS within 14 days in patients.

| Parameter | Favipiravir<br>(n = 62) |  | Control<br>(n = 31) |  | All<br>(n = 93) |  |
| --- | --- | --- | --- | --- | --- | --- |
|  | Adjusted<br>Odd ratio<br>(Robust<br>standard<br>errors) | P-value | Adjusted<br>Odd ratio<br>(Robust<br>standard<br>errors) | P-value | Adjusted<br>Odd ratio<br>(Robust<br>standard<br>errors) | P-value |
| Treatment |  |  |  |  |  |  |
| Non-favipiravir |  |  |  |  | Reference |  |
| Favipiravir |  |  |  |  | 9.92 (5.62) | < 0.001 |
| Age (continuous) | 0.95 (0.04) | 0.1614 | 0.99 (0.05) | 0.992 | 0.98 (0.03) | 0.453 |
| Gender |  |  |  |  |  |  |
| Male | Reference |  |  |  |  |  |
| Female | 6.35 (4.70) | < 0.001 | 0.69 (0.72) | 0.723 | 1.90 (1.12) | 0.275 |
| BMI |  |  |  |  |  |  |
| Normal weight | Reference |  |  |  |  |  |
| Underweight | 0.29 (0.27) | 0.180 | 3.87 (4.55) | 0.250 | 1.13 (0.90) | 0.878 |
| Overweight and<br>obese | 6.36 (8.88) | 0.185 | 1.01 (1.62) | 0.999 | 2.45 (1.70) | 0.197 |
| Viral genotype |  |  |  |  |  |  |

| Parameter | Favipiravir<br>(n = 62) |  | Control<br>(n = 31) |  | All<br>(n = 93) |  |
| --- | --- | --- | --- | --- | --- | --- |
|  | Adjusted<br>Odd ratio<br>(Robust<br>standard<br>errors) | P-value | Adjusted<br>Odd ratio<br>(Robust<br>standard<br>errors) | P-value | Adjusted<br>Odd ratio<br>(Robust<br>standard<br>errors) | P-value |
| Ancestral strain with<br>D614G | *Reference* |  |  |  |  |  |
| Alpha and Delta | 1.49 (1.26) | 0.639 | 0.45 (0.51) | 0.482 | 1.45 (0.87) | 0.536 |
| **Duration of symptoms**<br>**before treatment**<br>*(continuous)* | 1.23 (0.24) | 0.248 | 0.84 (0.21) | 0.497 | 1.10 (1.13) | 0.402 |
| **Have underlying health conditions** |  |  |  |  |  |  |
| No | *Reference* |  |  |  |  |  |
| Yes | 0.65 (0.75) | 0.939 | - |  | 0.55 (0.45) | 0.463 |
| **Constant** | **0.64 (0.75)** | **0.706** | **0.77 (1.39)** | **0.887** | **0.42 (0.39)** | **0.353** |

There was no significant difference in saliva viral load during treatment between the two arms (Figure 3A). However, saliva viral load levels were lower in the FPV arm on days 1 and 13 of treatment for participants with baseline viral loads in the lowest quartile, and on day 28 for those with baseline viral loads in the highest quartile (Figure 3 B, C). There was no significant difference in time to undetectable virus in saliva samples from FPV and control arms (median 19 vs 16 days; IQR of 10-28 days for both, aHR 0.96, 95% CI 0.58-1.58, *P* = 0.871, see Supplementary Figure 2). There was no correlation between time to sustained clinical improvement and time to undetectable virus in saliva (r = 0.13, *P* = 0.65).

**Figure 3.**
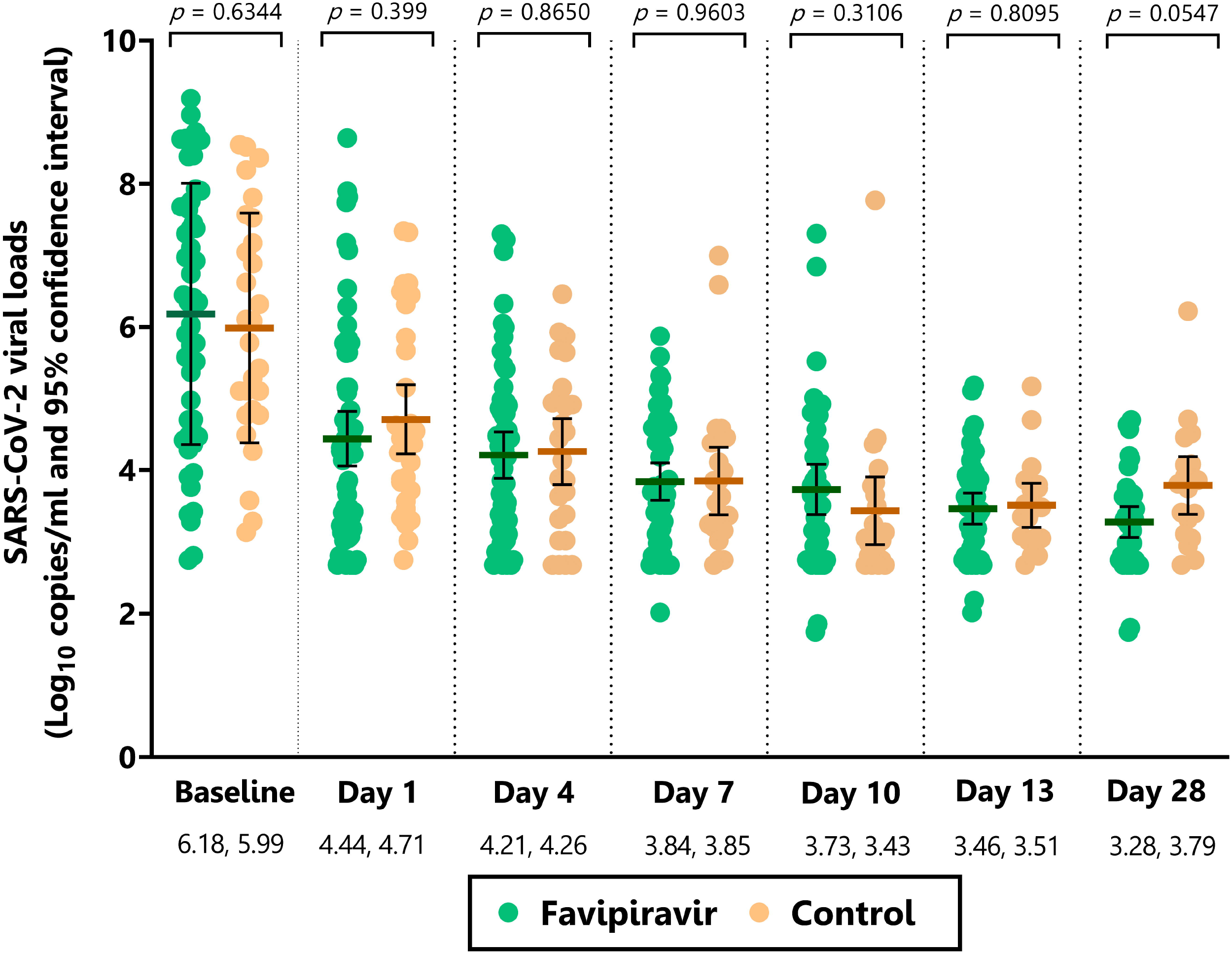
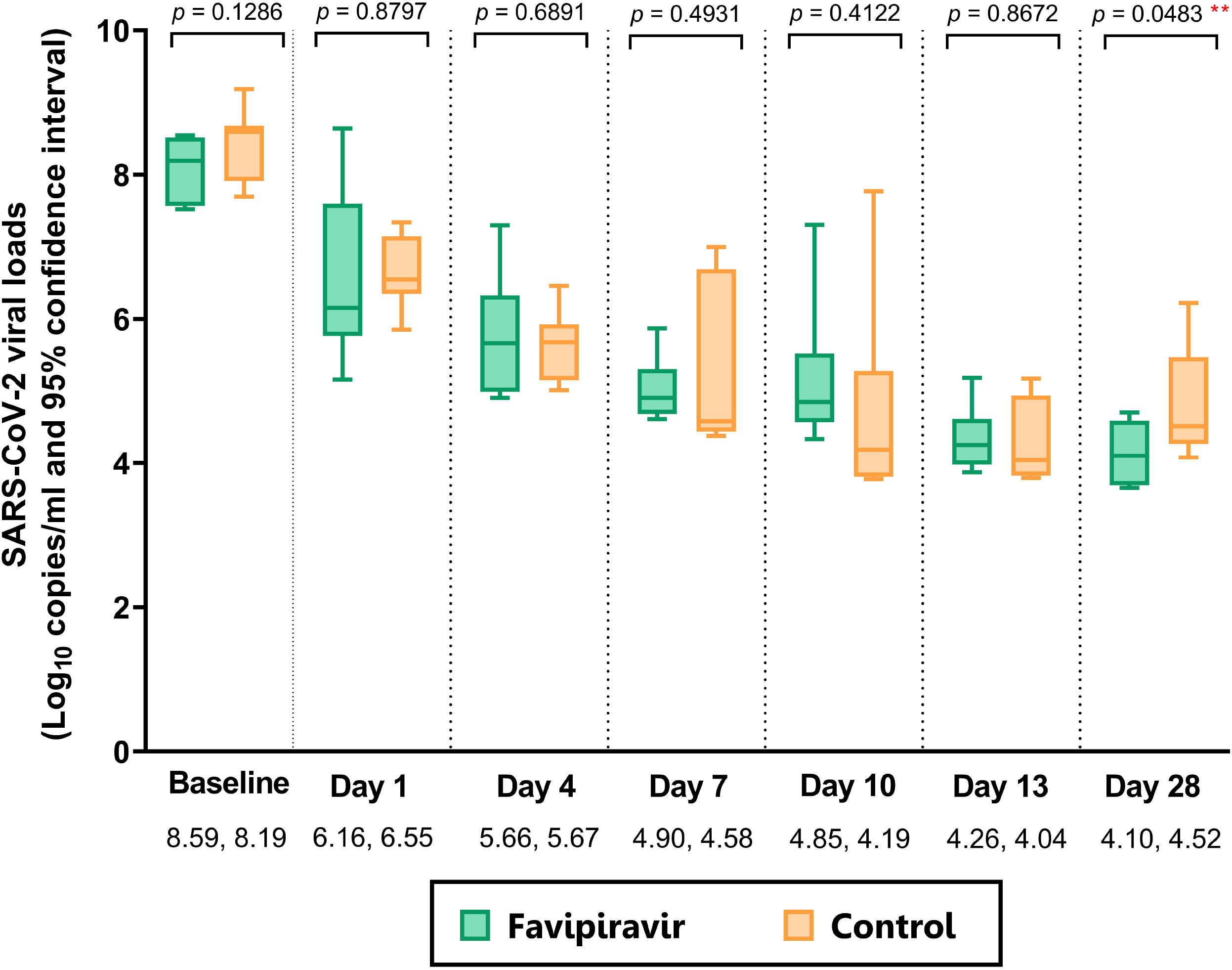
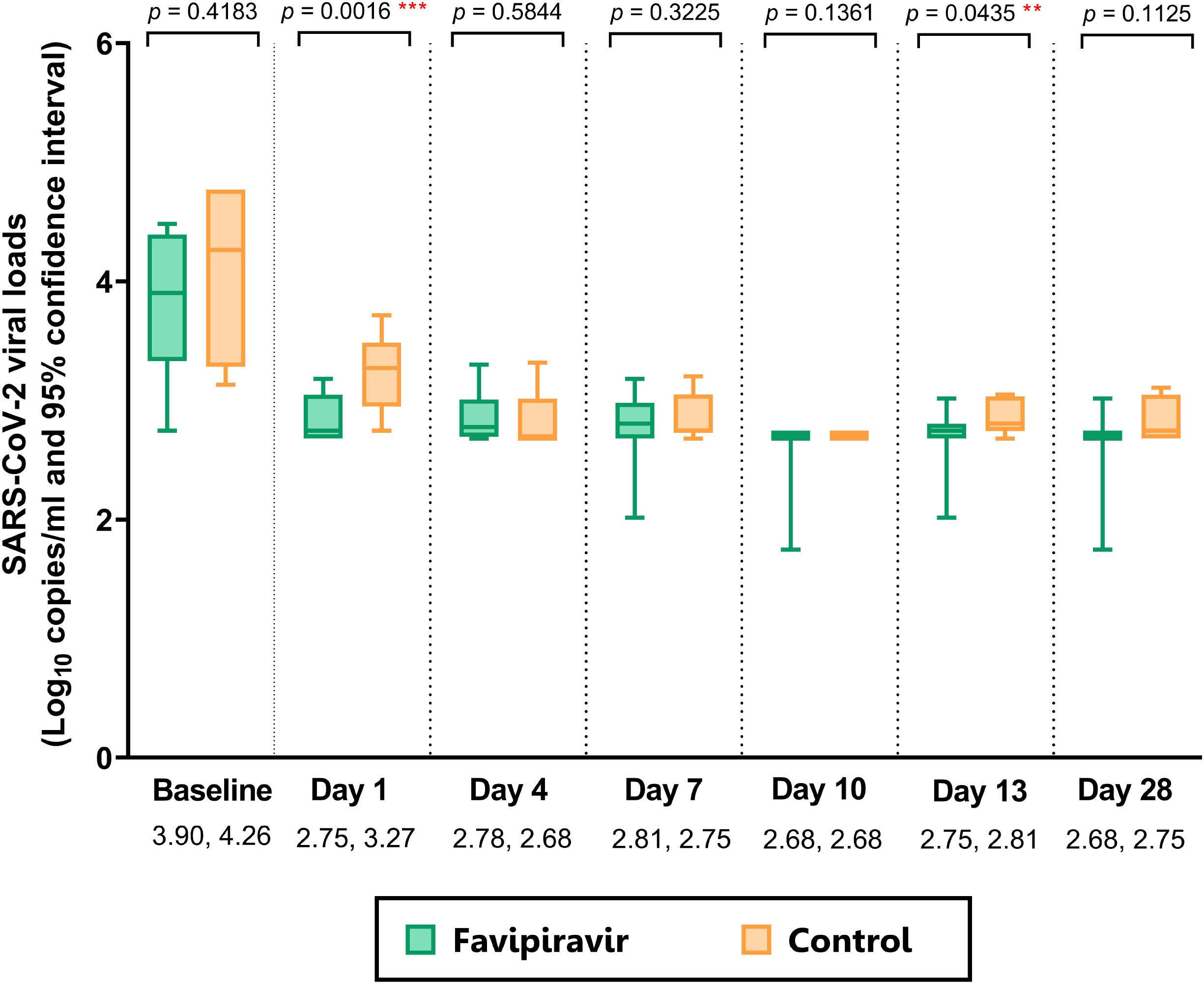
Quantitative SARS-CoV-2 viral loads over 0-28 days in FPV and control arms: (A) overall (B) participants with baseline viral load ≥75^th^ percentile and (C) participants with baseline viral load ≤25^th^ percentile.

Although the FPV arm showed significantly higher blood uric acid levels on days 4, 7, and 10 (*P* < 0.001 for all time points, see Supplementary Figure 3 and Supplementary Table 1), there were no associated clinical symptoms and blood uric acid levels became normalized by day 28, with no difference between arms. Participants in the FPV arm had higher alanine aminotransferase (ALT) levels on day 10 (mean (range) of 32.0 (25.9-40.1) U/L and 26.6 (24.5-31.6) U/L for FPV and control arms respectively, where *P* = 0.0258), and longer QT intervals on days 14 and 28 (*P* < 0.001 for both days). Although, all values were within normal limits (see Supplementary Figure 3 and Supplementary Table 1). There were no differences in C-reactive protein and procalcitonin levels during treatment between arms.

Thirty-six and 10 adverse events (AEs) were respectively reported FPV and control arms, all relatively mild and fully resolved within the 28^th^ day of treatment. FPV-related AEs, as determined by investigator, were: hyperuricemia (n = 11), maculopapular rashes (n = 3), leukopenia (n = 1), and increased SGPT (n = 1). One of the three patients that developed a maculopapular rash discontinued FPV treatment. All three cases were relatively mild, and completely resolved by the end of the study. 8 (12.9%) and 7 (22.6%) patients in FPV and control arms (*P* = 0.316) respectively developed mild pneumonia at a median (range) 6.5 (1-13) and 7 (1-13) days after treatment, respectively; all recovered well without complications.

## Discussion

This prospective cohort evaluated the clinical efficacy of FPV treatment at a dosage of 1800/800 BID for 5-14 days in patients with mild-to-moderate COVID-19 without pneumonia. Patients received treatment for an average 1.6 days after disease onset (90% before 4 days). Using the unbiased NEWS clinical severity assessment system, we found that patients (females in particular) treated with FPV were significantly more likely to experience clinical improvement from COVID-19 within 14 days than controls. FPV-administered patients had shorter time to sustained clinical improvement – a median 2 days compared to 14 days without FPV. There was no evident benefit on overall viral load, although patients with lower baseline viral loads had greater viral reductions on days 1 and 13 of treatment. Patients in the FPV arm were less likely to develop pneumonia, although this was not statistically significant. FPV was generally well tolerated but often associated with asymptomatic hyperuricemia.

These findings coincide with multiple reviews and meta-analyses supporting FPV’s clinical efficacy after 7–14-day regimens [13,14,25,32,33,50]. The Japanese Association for Infectious Diseases reported rates of symptomatic improvement after a 14-day course of FPV in 90% of patients with mild COVID-19 [28]. Sawanpanyalert et al (2021) demonstrated that the 140 Thai patients admitted and treated with FPV within 4 days upon symptomatic onset, in conjunction to other treatment modalities, had significantly lower odds of experiencing poor outcomes compared to those initiated treatment after 4 days of onset. Similarly, other studies also showed that early treatment, particularly by 4 days of onset, was associated with earlier defervescence [8,45]. These reports highlight the importance of early intervention and its clinical benefits, as observed in our study.

Many studies also demonstrated FPV’s ability to inhibit viral progression and promote viral clearance compared to other antivirals [4,13,51,52]. Doi et al (2020) emphasized that more efficient and rapid viral clearance rates and defervescence were respectively observed upon early treatment, and Ivashchenko et al (2021) discussed further how prolonged treatment regimens promoted this clearance. A recent meta-analysis revealed that FPV treatment in mild-to-moderate cases of COVID-19 was associated with higher viral clearance and shorter hospital stay, but not beneficial in severe COVID-19 cases [52]. Other studies revealed lack of benefit of FPV treatment due to insufficient evidence that FPV affects rates of mortality, mechanical ventilation, and viral clearance [31-33]. Our study with mild-to-moderate COVID-19 patients also found no significant difference in viral clearance rate, despite clear clinical improvement – except in the subgroup with low baseline viral load. However, viral clearance may not be an appropriate measure of treatment efficacy, as some patients may have recovered – or even be symptom-free – but still have detectable high viral titres [31]. Furthermore, drugs with a lethal mutagenesis mechanism of action may be more prone to the inadequacies of a viral RNA endpoint because viral RNA may theoretically be sufficiently conserved within the primer/probe target sequences so as to be detected, but mutated elsewhere to a degree that they are unable to produce infectious virus. Indeed, while an impact upon viral RNA clearance was detected in phase II for MPV (which also executes lethal mutagenesis), more marked differences in infectious virus titres were observed [53].

While demonstrating significant clinical benefit, RDV treatment in 562 non-hospitalized patients showed no efficacy on viral clearance [19]. As late-phase inflammatory responses lie behind severe COVID-19 illness, antiviral treatment may only prove beneficial towards controlling viral replication upon early administration, therefore reducing subsequent inflammatory responses. This could explain the limited clinical benefits of RDV observed in the WHO Solidarity Trial Consortium (2021) and DisCoVeRy trials [54] across patients of different clinical conditions and varied treatment initiation.

Other studies also found FPV to be safe for short-term usage, with relatively mild or moderate AEs [52]. Some typically observed side effects included: hyperuricemia; elevated triglycerides, serum ALT, and serum uric acid; gastrointestinal discomfort; and abnormal liver function [8,9,11,13,33]. No significant differences in incidence and prevalence of AEs between FPV and control arms were observed [32,33,50,52]. These events are thought to be related to elevated liver function, QT prolongation, skin rashes, and OAT1, OAT3, and URAT1 receptor inhibition [28]. The FPV regimen in our study appeared well tolerated by participants in general. A majority experienced relatively mild AEs and all fully recovered thereafter.

Our study has some clear limitations. The first was its open label, which could lead to subjective symptomatology bias. We avoided using self-reported symptoms and used NEWS to quantify clinical response instead. We did not assess improvement of subjective symptoms (*e*.*g*., cough, sore throat, headache, nasal congestion), which could be affected by other factors. The second limitation was that our results cannot be directly applied to severe cases of COVID-19, as we targeted mild-to-moderate COVID-19 cases without pneumonia. Most of the participants were relatively healthy, making extrapolations to patient at risk of severe COVID-19 difficult – as found in another study [55]. The third limitation was our small sample size, which made it difficult to demonstrate the benefits of preventing disease progression. Although, we did observe a decreased prevalence of pneumonia in the FPV arm.

To summarize, our findings support previous literature that early administration of FPV in mild COVID-19 expedites recovery, and is relatively safe for short-term usage. This is relevant to a majority of patients, who are affected by mild SARS-CoV-2. While novel antiviral agents *(e*.*g*., nirmatrelvir/ritonavir and MPV) continue to emerge and substantive efforts are underway to address access for low-and middle-income countries [56,57], their current exorbitant prices, restricted accessibility in resource-limited settings, and poorly characterized pharmacokinetic and safety profiles would not render them a first choice in resource-limited settings for the time being. Publicly available information regarding the pharmacokinetics of FPV in different populations are currently extremely sparse, but will be critical to understand whether some of the differences between studies can be explained by regional differences in exposure. More data is required in this area, but current understanding does support that antiviral concentrations may be achieved within doses already administered to humans [58]. Due to its safety and comparative current cost, FPV may be a suitable treatment for mild COVID-19 without pneumonia. Further studies are required to evaluate the benefits of FPV treatment on post-acute COVID-19 syndrome and other complications.

## Data Availability

All data produced in the present study are available upon reasonable request to the authors

## Acknowledgements

This study was developed in collaboration of Faculty of Medicine Siriraj Hospital, Mahidol University and National Centre for Global Health and Medicine, Japan. The study drug (favipiravir) was supported by FUJIFILM Toyama Chemical Co., Ltd. We would like to thank all the staff in the three participating hospitals who provided support for this study.

