## Supplementary Table 1-3, Supplementary Figure 1-3 for "Early Treatment of Favipiravir in COVID-19 Patients Without Pneumonia: A Multicentre, Open-Labelled, Randomized Control Study"

Supplementary Table 1. Clinical laboratory evaluations of SARS-CoV-2 infected patients 0-28 days after infection.

| Clinical Laboratory Tests | Mean (SE) | | P-value |
|---|---|---|---|
| | Case | Control | |
| **(1) Complete Blood Count (CBC)** | | | |
| **1.1) White Blood Cells (WBC) count** *(Reference range: female 4.5-8.0 x $10^3$/µL, male4.5-9.0 x $10^3$/µL)* | | | |
| Day 0 (Screening days) | 5.24 (2.23) | 5.15 (0.30) | 0.8210 |
| Day 4 | 5.14 (0.19) | 6.45 (1.03) | 0.0939 |
| Day 7 | 7.17 (0.97) | 6.15 (0.34) | 0.4680 |
| Day 10 | 5.84 (0.22) | 7.00 (0.38) | <0.001 |
| Day 28 | 6.45 (0.19) | 6.68 (0.31) | 0.5047 |
| **1.2) Red Blood Cell (RBC) count** *(Reference range: female 3.7-5.0 x $10^6$/uL, male 4.0-5.7* | | | |
| Day 0 (Screening days) | 4.86 (0.08) | 4.88 (0.20) | 0.8760 |
| Day 4 | 4.80 (0.14) | 5.05 (0.13) | 0.2409 |
| Day 7 | 4.89 (0.07) | 5.08 (0.12) | 0.1721 |
| Day 10 | 4.79 (0.30) | 5.06 (0.29) | 0.1638 |
| Day 28 | 4.56 (0.10) | 4.67 (0.20) | 0.5813 |
| **1.3) Haemoglobin (Hb)** *(Reference range: female 11.0-14.0 g/dL, male 12.0-15.0 g/dL)* | | | |
| Day 0 (Screening days) | 13.13 (0.32) | 13.56 (0.30) | 0.3970 |
| Day 4 | 13.51 (0.29) | 13.73 (0.31) | 0.6307 |
| Day 7 | 13.31 (0.26) | 13.61 (1.63) | 0.4864 |

| Clinical Laboratory Tests | Mean (SE) | | P-value |
|---|---|---|---|
| | Case | Control | |
| Day 10 | 13.22 (0.30) | 13.66 (0.29) | 0.3354 |
| Day 28 | 12.53 (0.24) | 12.73 (0.26) | 0.5968 |
| **1.4) Haematocrit (Hct)** *(Reference range: female 35.0-41.0 %, male 38.0-48.0 %)* | | | |
| Day 0 (Screening days) | 40.04 (0.80) | 40.96 (0.82) | 0.4711 |
| Day 4 | 40.42 (0.79) | 41.33 (0.84) | 0.4737 |
| Day 7 | 39.84 (0.71) | 41.08 (0.77) | 0.2782 |
| Day 10 | 46.72 (7.39) | 40.81 (0.76) | 0.5540 |
| Day 15 | 39.54 (0.97) | 40.21 (1.71) | 0.7275 |
| Day 22 | 40.80 (0.97) | 37.35 (0.35) | 0.1129 |
| Day 28 | 38.24 (0.63) | 38.57 (0.66) | 0.7469 |
| **(2) Haematology Laboratory Normal Range (count.)** | | | |
| **2.1) Platelet** *(Reference range: 100-400 x $10^3$/uL)* | | | |
| Day 0 (Screening days) | 243.25 (10.76) | 246.35 (10.58) | 0.8553 |
| Day 4 | 248.39 (12.75) | 237.07 (10.85) | 0.5725 |
| Day 7 | 275.28 (11.72) | 257.13 (11.81) | 0.3313 |
| Day 10 | 284.34 (11.59) | 277.05 (16.51) | 0.7140 |
| Day 28 | 293.67 (9.89) | 258.76 (12.82) | 0.0406 |
| **(3) Differential WBC count** | | | |
| **3.1) Neutrophil** *(Reference range: female 35-75 %, male 36-70 %)* | | | |
| Day 0 (Screening days) | 56.21 (1.82) | 54.87 (2.07) | 0.6534 |

| Clinical Laboratory Tests | Mean (SE) | | P-value |
|---|---|---|---|
| | Case | Control | |
| Day 4 | 52.40 (1.30) | 52.19 (1.41) | 0.9213 |
| Day 7 | 55.05 (1.46) | 52.06 (2.24) | 0.2520 |
| Day 10 | 58.32 (1.62) | 55.62 (2.19) | 0.3233 |
| Day 28 | 55.77 (0.82) | 55.34 (1.76) | 0.7990 |
| **3.2) Lymphocyte** *(Reference range: female 20-59 %, male 23-57 %)* | | | |
| Day 0 (Screening days) | 32.62 (1.45) | 35.17 (1.98) | 0.3062 |
| Day 4 | 36.63 (1.14) | 37.69 (1.47) | 0.5842 |
| Day 7 | 34.75 (1.24) | 38.56 (1.93) | 0.0893 |
| Day 10 | 31.20 (1.43) | 35.14 (1.96) | 0.1068 |
| Day 28 | 33.29 (0.83) | 33.89 (1.65) | 0.7171 |
| **3.3) Monocyte** *(Reference range: 2-10 %)* | | | |
| Day 0 (Screening days) | 8.61 (0.62) | 8.10 (0.45) | 0.5844 |
| Day 4 | 7.67 (0.38) | 6.93 (0.57) | 0.2709 |
| Day 7 | 7.26 (0.41) | 6.20 (0.43) | 0.1099 |
| Day 10 | 7.11 (0.25) | 5.72 (0.34) | <0.001 |
| Day 28 | 7.25 (0.22) | 6.48 (0.28) | 0.038 |
| **3.4) Eosinophil** *(Reference range: 1-5 %)* | | | |
| Day 0 (Screening days) | 1.86 (0.27) | 1.45 (0.36) | 0.3814 |
| Day 4 | 2.55 (0.26) | 2.61 (0.44) | 0.8947 |
| Day 7 | 2.34 (0.27) | 2.38 (0.37) | 0.9292 |
| Day 10 | 2.62 (0.24) | 2.97 (0.49) | 0.4746 |

| Clinical Laboratory Tests | Mean (SE) | | P-value |
|---|---|---|---|
| | Case | Control | |
| Day 28 | 3.44 (0.33) | 3.61 (0.45) | 0.7614 |
| **3.5) Basophil** *(Reference range: 0-3 %)* | | | |
| Day 0 (Screening days) | 0.27 (0.06) | 0.22 (0.07) | 0.6211 |
| Day 4 | 0.29 (0.06) | 0.28 (0.08) | 0.9191 |
| Day 7 | 0.39 (0.11) | 0.91 (0.66) | 0.2951 |
| Day 10 | 0.46 (0.07) | 0.38 (0.09) | 0.4644 |
| Day 28 | 0.54 (0.07) | 0.51 (0.90) | 0.8621 |
| **(4) Clinical Chemistry** | | | |
| **4.1) Albumin** *(Reference range: 3.5-5.0 g/dL)* | | | |
| Day 0 (Screening days) | 4.21 (0.07) | 4.26 (0.06) | 0.6233 |
| Day 4 | 3.88 (0.09) | 4.14 (0.51) | 0.0500 |
| Day 7 | 4.02 (0.04) | 4.11 (0.04) | 0.1578 |
| Day 10 | 3.97 (0.04) | 4.10 (0.06) | 0.0633 |
| Day 28 | 4.19 (0.04) | 4.24 (0.05) | 0.4728 |
| **4.2) BUN** *(Reference range: 6-20 mg/dL)* | | | |
| Day 0 (Screening days) | 10.32 (0.43) | 10.06 (0.44) | 0.7147 |
| Day 4 | 10.50 (0.28) | 10.33 (0.35) | 0.7208 |
| Day 7 | 11.21 (0.42) | 10.77 (0.50) | 0.5205 |
| Day 10 | 10.85 (0.46) | 11.07 (0.59) | 0.7719 |
| Day 28 | 11.97 (0.51) | 11.11 (0.69) | 0.3332 |

| Clinical Laboratory Tests | Mean (SE) | | P-value |
|---|---|---|---|
| | Case | Control | |
| **4.3) Alanine aminotransferase: ALT (SGPT)** *(Reference range: female 0-31 U/L, male 0-41 U/L)* | | | |
| Day 0 (Screening days) | 25.89 (2.36) | 27.48 (6.09) | 0.7694 |
| Day 4 | 27.28 (2.31) | 24.76 (5.25) | 0.6104 |
| Day 7 | 31.10 (2.61) | 24.63 (3.81) | 0.1601 |
| Day 10 | 36.06 (3.41) | 24.55 (2.92) | 0.0258 |
| Day 28 | 40.02 (4.73) | 31.63 (8.94) | 0.3823 |
| **4.3) Aspartate aminotransferase: AST (SGOT)** *(Reference range: female 0-31 U/L, male 0-37 U/L)* | | | |
| Day 0 (Screening days) | 28.33 (2.23) | 27.87 (2.63) | 0.9002 |
| Day 4 | 26.70 (1.87) | 23.76 (1.87) | 0.3302 |
| Day 7 | 28.65 (2.18) | 22.33 (1.89) | 0.0642 |
| Day 10 | 28.22 (2.12) | 22.62 (1.18) | 0.0636 |
| Day 28 | 26.64 (1.62) | 24.98 (3.02) | 0.5964 |
| **4.4) Alkaline phosphatase: ALP** *(Reference range: 35-129 U/L)* | | | |
| Day 0 (Screening days) | 63.49 (2.43) | 64.34 (7.46) | 0.8910 |
| Day 4 | 66.92 (2.23) | 59.97 (3.05) | 0.0746 |
| Day 7 | 72.01 (3.34) | 63.93 (3.36) | 0.1304 |
| Day 10 | 69.42 (2.82) | 64.90 (4.25) | 0.3614 |
| Day 28 | 68.22 (2.29) | 60.78 (3.74) | 0.0798 |
| **4.5) Calcium** *(Reference range: 8.6-10 mg/dL)* | | | |

| Clinical Laboratory Tests | Mean (SE) | | P-value |
|---|---|---|---|
| | Case | Control | |
| Day 0 (Screening days) | 9.20 (0.05) | 9.29 (0.08) | 0.3417 |
| Day 4 | 9.14 (0.04) | 9.32 (0.72) | 0.0301 |
| Day 7 | 9.29 (0.05) | 9.38 (0.08) | 0.3333 |
| Day 10 | 9.60 (0.29) | 9.53 (0.09) | 0.8684 |
| Day 28 | 9.33 (0.05) | 11.14 (1.70) | 0.1303 |
| **4.6) Phosphorus** *(Reference range: 2.7-4.5 mg/dL)* | | | |
| Day 0 (Screening days) | 3.44 (0.09) | 3.24 (0.09) | 0.1548 |
| Day 4 | 3.55 (0.08) | 3.58 (0.08) | 0.8544 |
| Day 7 | 3.58 (0.70) | 3.60 (0.97) | 0.9550 |
| Day 10 | 4.62 (0.81) | 3.80 (0.09) | 0.4434 |
| Day 28 | 3.43 (0.07) | 3.52 (0.23) | 0.6453 |
| **4.7) Potassium** *(Reference range: 3.5-5.1 mmol/L)* | | | |
| Day 0 (Screening days) | 3.83 (0.08) | 3.73 (0.04) | 0.3766 |
| Day 4 | 3.91 (0.09) | 3.28 (0.05) | 0.5162 |
| Day 7 | 4.53 (0.65) | 3.81 (0.06) | 0.4358 |
| Day 10 | 3.93 (0.05) | 5.16 (1.28) | 0.2022 |
| Day 28 | 5.96 (2.26) | 4.56 (0.84) | 0.6737 |
| **4.8) Sodium** *(Reference range: 136-145 mmol/L)* | | | |
| Day 0 (Screening days) | 136.24 (2.02) | 138.16 (0.50) | 0.5083 |
| Day 4 | 137.97 (0.26) | 138.23 (0.32) | 0.5414 |
| Day 7 | 136.77 (2.09) | 138.77 (0.39) | 0.5040 |

| Clinical Laboratory Tests | Mean (SE) | | P-value |
|---|---|---|---|
| | Case | Control | |
| Day 10 | 137.21 (2.44) | 139.24 (0.49) | 0.5385 |
| Day 28 | 136.51 (2.68) | 128.92 (6.57) | 0.1786 |
| **4.9) Chloride** *(Reference range: 98-107 mmol/L)* | | | |
| Day 0 (Screening days) | 101.92 (1.65) | 102.94 (0.39) | 0.6628 |
| Day 4 | 103.25 (0.29) | 99.17 (3.25) | 0.0965 |
| Day 7 | 103.57 (0.34) | 104.23 (1.14) | 0.4791 |
| Day 10 | 102.54 (1.84) | 102.93 (0.41) | 0.8767 |
| Day 28 | 103.3 (0.27) | 100.91 (3.48) | 0.3294 |
| **4.10) Uric acid** *(Reference range: female 2.4-5.7 mg/dL, male 3.4-7.0 mg/dL)* | | | |
| Day 0 (Screening days) | 5.03 (0.18) | 5.30 (0.29) | 0.4071 |
| Day 4 | 8.68 (0.18) | 5.10 (0.22) | <0.001 |
| Day 7 | 9.08 (0.25) | 5.26 (0.31) | <0.001 |
| Day 10 | 8.93 (0.29) | 5.74 (0.35) | <0.001 |
| Day 28 | 5.58 (0.18) | 5.92 (0.46) | 0.431 |
| **(5) Clinical Immunology Laboratory Normal Range (Cont.)** | | | |
| **5.1) C-Reactive Protein (CRP)** *(Reference range: 0-1 ug/dL)* | | | |
| Day 0 (Screening days) | 0.77 (0.14) | 1.26 (0.47) | 0.2080 |
| Day 4 | 0.85 (0.19) | 0.513 (0.19) | 0.2714 |
| Day 7 | 0.79 (0.16) | 0.81 (0.23) | 0.9576 |
| Day 10 | 0.52 (0.11) | 0.77 (0.26) | 0.3099 |
| Day 28 | 0.55 (0.13) | 0.75 (0.20) | 0.3880 |

| Clinical Laboratory Tests | Mean (SE) | | P-value |
|---|---|---|---|
| | Case | Control | |
| **5.2) Procalcitonin (PCT)** *(Reference range: 0- 0.5 ng/dL)* | | | |
| Day 0 (Screening days) | 0.46 (0.12) | 0.56 (0.31) | 0.7158 |
| Day 4 | 0.47 (0.15) | 0.61 (0.31) | 0.6245 |
| Day 7 | 0.52 (0.19) | 0.33 (0.16) | 0.5379 |
| Day 10 | 0.34 (0.14) | 0.36 (0.18) | 0.9490 |
| Day 28 | 0.59 (0.16) | 0.49 (0.22) | 0.7170 |
| **(6) Cardiovascular Lab** | | | |
| **6.1) QT interval** *(Reference range: female < 470 ms, male < 450 ms)* | | | |
| Day 0 (Screening days) | 378.32 (5.91) | 379.63 (3.79) | 0.8834 |
| Day 14 | 427.45 (5.81) | 401.68 (6.63) | <0.001 |
| Day 28 | 392.60 (3.19) | 378.72 (4.93) | <0.001 |
| **6.2) PR interval** *(Reference range: 120-200 ms)* | | | |
| Day 0 (Screening days) | 156.66 (8.10) | 144.07 (3.44) | 0.2904 |
| Day 14 | 153.49 (4.55) | 147.89 (5.38) | 0.4612 |
| Day 28 | 152.68 (2.81) | 149.17 (4.79) | 0.5042 |

Supplementary Table 2. Prevalence of adverse events by group.

| Adverse Event | Favipiravir (n=62) (%) | Control (n=31) (%) |
|---|---|---|
| Any AE (e.g., unspecified fever, upper respiratory tract infection, acute lymphadenitis right cervical, steroid acne, etc.) | 10 (16.1) | 2 (6.5) |
| Constipation | 2 (3.2) | - |
| Diarrhoea | 1 (1.6) | - |
| Dizziness | 1 (1.6) | - |
| Dyspepsia | 1 (1.6) | - |
| Insomnia | 2 (3.2) | - |
| Rash | 3 (4.8) | - |
| Hypertension | - | 1 (3.2) |
| Hypertriglyceridemia | 2 (3.2) | - |
| Hyperuricemia | 11 (17.7) | 1 (3.2) |
| Hypokalaemia | 6 (9.7) | 7 (22.6) |
| Hypoleukaemia | 1 (1.6) | - |
| Increased CPK | - | 1 (3.2) |
| Increased SGPT | 1 (1.6) | - |
| Anaemia | 2 (3.2) | - |

| Adverse Event | Favipiravir (n=62) (%) | Control (n=31) (%) |
|---|---|---|
| Leukopenia | 1 (1.6) | - |
| Hyperglycaemia | 2 (3.2) | - |

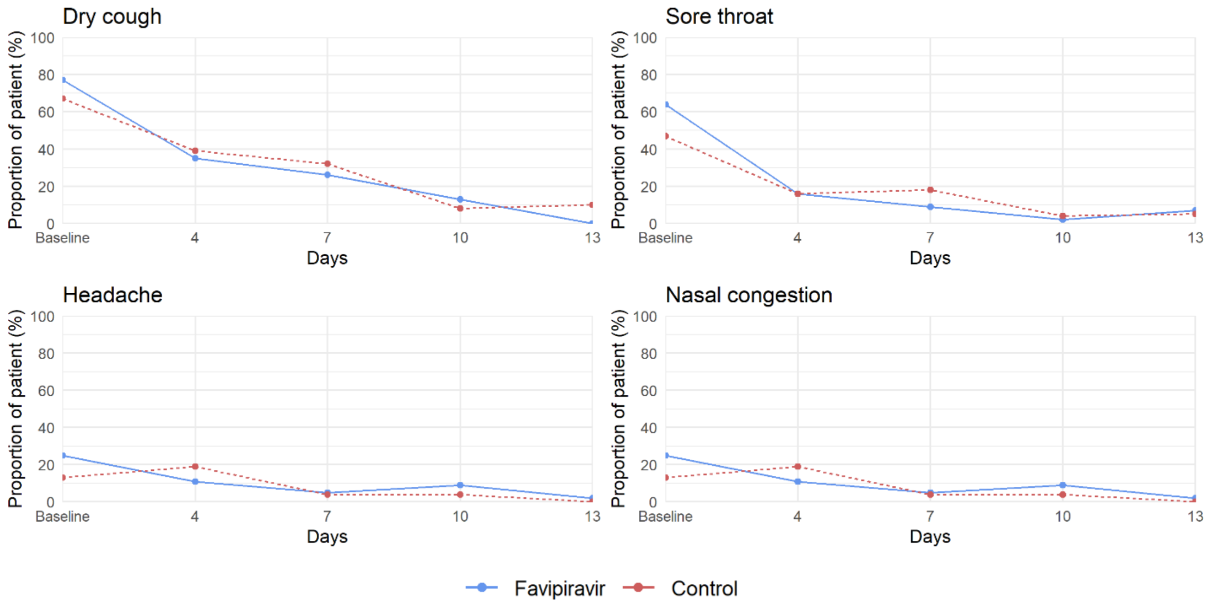


Supplementary Figure 1. Changes in clinical symptoms (dry cough, sore throat, headache, and nasal congestion) over 0-13 days. Line graphs illustrate the proportion of patients who had clinical symptoms each day after enrolment.

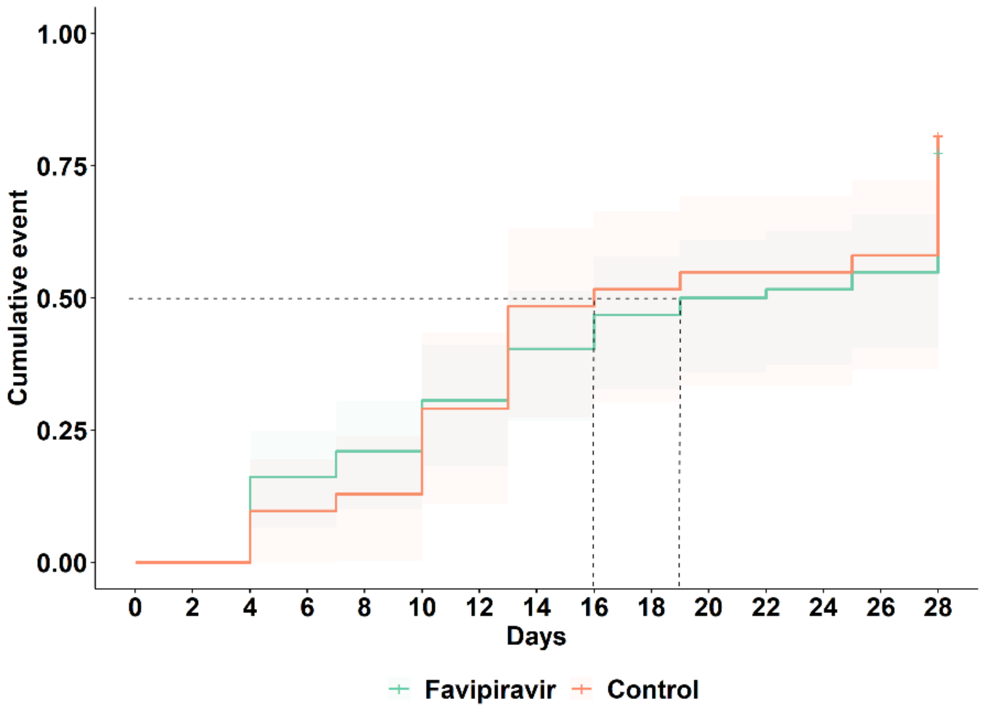


Supplementary Figure 2. Time to undetectable virus over 28 days. The Kaplan-Meier curve illustrates the cumulative proportion of patients who had undetected viral titres over 28 days. The median time to undetected virus was 19 days vs 16 days (interquartile range (IQR) of 10-28 days for both) for FPV and control arms respectively (adjusted hazard ratio (aHR) 0.96, 95% CI: 0.58-1.58, *P* = 0.871).

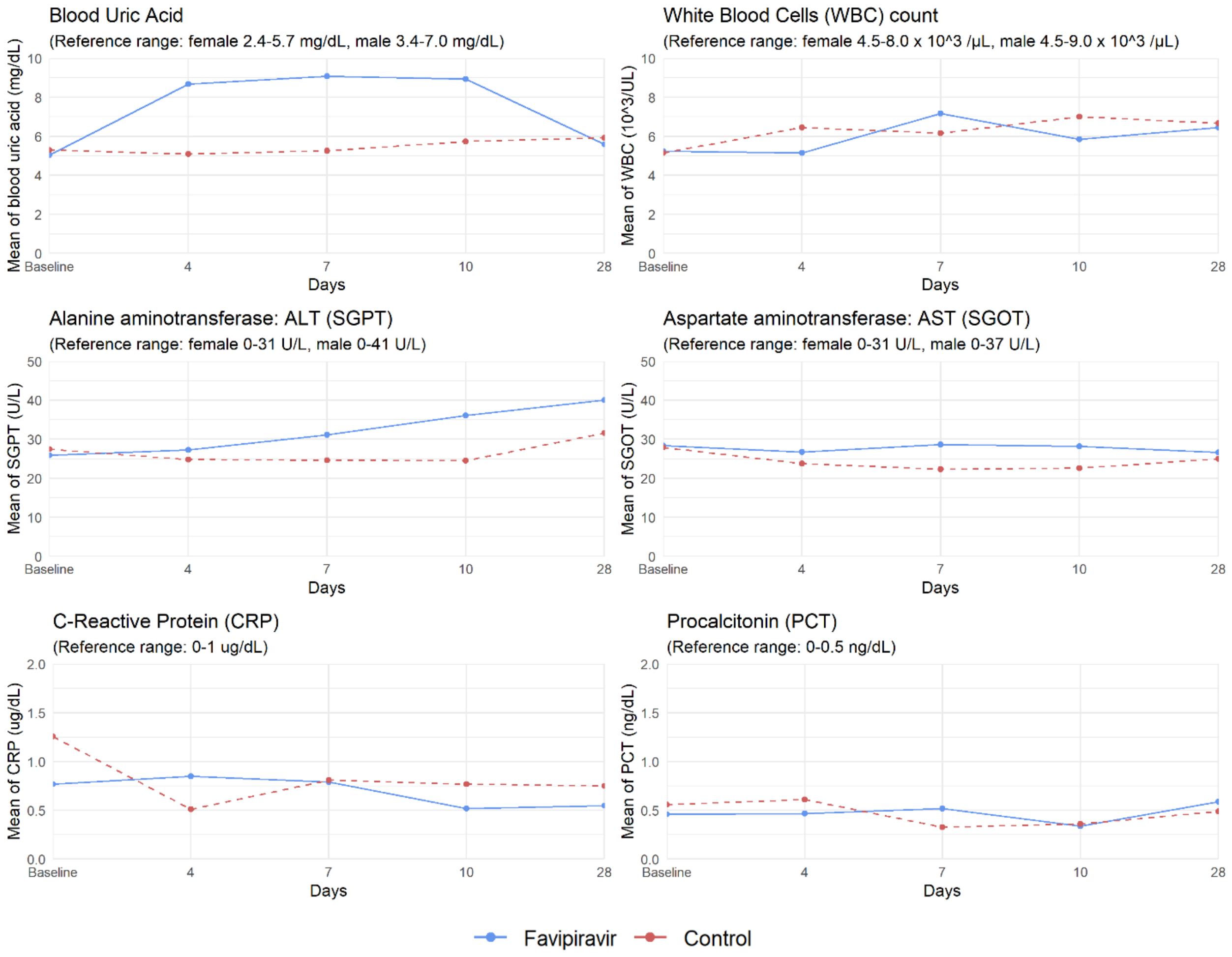


Supplementary Figure 3. Clinical laboratory evaluations of blood uric acid, white blood cell (WBC) count, alanine aminotransferase (ALT), aspartate aminotransferase (AST), C-reactive protein (CRP), and procalcitonin (PCT). Line graphs illustrate the proportion of patients who had clinical symptoms from days 0-28.
